# Development and validation of the COVID-19 severity index (CSI): a prognostic tool for early respiratory decompensation

**DOI:** 10.1101/2020.05.07.20094573

**Authors:** Adrian Haimovich, Neal G. Ravindra, Stoytcho Stoytchev, H. Patrick Young, Francis Perry Wilson, David van Dijk, Wade L. Schulz, R. Andrew Taylor

**Affiliations:** Department of Emergency Medicine, Yale University School of Medicine, New Haven, CT; Department of Internal Medicine, Section of Cardiovascular Medicine, Yale University School of Medicine, New Haven, CT; Department of Computer Science, Yale University, New Haven, CT; Center for Outcomes Research and Evaluation, Yale New Haven Hospital, New Haven, CT; Department of Internal Medicine, Yale University School of Medicine, New Haven, CT; Clinical and Translational Research Accelerator, Department of Medicine, Yale University School of Medicine, New Haven CT; Center for Medical Informatics, Yale University School of Medicine, New Haven, CT; Department of Laboratory Medicine, Yale University School of Medicine, New Haven, CT

## Abstract

**Objective:** The goal of this study was to create a predictive model of early hospital respiratory decompensation among patients with COVID-19.

**Design:** Observational, retrospective cohort study.

**Setting:** Nine-hospital health system within the Northeastern United States.

**Populations:** Adult patients (≥ 18 years) admitted from the emergency department who tested positive for SARS-CoV-2 (COVID-19) up to 24 hours after initial presentation. Patients meeting criteria for respiratory critical illness within 4 hours of arrival were excluded.

**Main outcome and performance measures:** We used a composite endpoint of critical illness as defined by oxygen requirement (greater than 10 L/min by low-flow device, high-flow device, non-invasive, or invasive ventilation) or death within the first 24 hours of hospitalization. We developed models predicting our composite endpoint using patient demographic and clinical data available within the first four hours of arrival. Eight hospitals (*n* = 932) were used for model development and one hospital (*n* = 240) was held out for external validation. Area under receiver operating characteristic (AU-ROC), precision-recall curves (AU-PRC), and calibration metrics were used to compare predictive models to three illness scoring systems: Elixhauser comorbidity index, qSOFA, and CURB-65.

**Results:** During the study period from March 1, 2020 to April 27,2020, 1,792 patients were admitted with COVID-19. Six-hundred and twenty patients were excluded based on age or critical illness within the first 4 hours, yielding 1,172 patients in the final cohort. Of these patients, 144 (12.3%) met the composite endpoint within the first 24 hours. We first developed a bedside quick COVID-19 severity index (qCSI), a twelve-point scale using nasal cannula flow rate, respiratory rate, and minimum documented pulse oximetry. We then created a machine-learning gradient boosting model, the COVID-19 severity index (CSI), using twelve additional variables including inflammatory markers and liver chemistries. Both the qCSI (AU-ROC mean [95% CI]: 0.90 [0.85-0.96]) and CSI (AU-ROC: 0.91 [0.86-0.97]) outperformed the comparator models (qSOFA: 0.76 [0.69-0.85]; Elixhauser: 0.70 [0.62-0.80]; CURB-65: AU-ROC 0.66 [0.58-0.77]) on cross-validation and performed well on external validation (qCSI: 0.82, CSI: 0.76, CURB-65: 0.50, qSOFA: 0.59, Elixhauser: 0.61). We find that a qCSI score of 0-3 is associated with a less than 5% risk of critical respiratory illness, while a score of 9-12 is associated with a 57% risk of progression to critical illness.

**Conclusions:** A significant proportion of admitted COVID-19 patients decompensate within 24 hours of hospital presentation and these events are accurately predicted using bedside respiratory exam findings within a simple scoring system.

## Introduction

The SARS-CoV-2 disease (COVID-19) is increasingly understood to be a disease with a significant rate of critical illness. International reports of intensive care unit (ICU) utilization frequencies have varied from less than 10% to above 30%.^1–3^ There are now reports from larger ICU cohorts, but these do not report a denominator of total COVID-19 population.^4,5^ More recently, a large New York City, USA case series was presented, of which 14.2% of patients with known outcomes were admitted to the ICU.^6^ Preliminary data from a second New York City, USA cohort had an ICU rate of 32.5%.^7^

While there is a growing body of data about critically ill cohorts and outcomes, less is known about risk factors for critical illness, especially as they relate to respiratory status. Oxygen saturation and inflammatory markers including d-dimer, ferritin, and C-reactive protein (CRP) have been identified as potentially associated with critical illness.^7^ Predictive models advance the purposes of risk factor analysis and, ideally, lay the groundwork for the assignment of individualized illness probabilities. A number of diagnostic and prognostic prediction models for COVID-19 have been proposed, but the included cohorts were small and at significant risk for bias.^8^

In this work, we expand on previous efforts describing the course of critical COVID-19 illness in three ways. First, we describe the prevalence of patient respiratory deterioration early (< 24 hours) during hospitalization. We focus on this time interval because of its significant implications both for resource utilization and for anticipatory guidance for patients and families. In many hospital systems, clinically deteriorating patients are evaluated urgently in consideration of higher levels of care—a process both personnel-intensive often including ward providers, a rapid response team, and critical care consultants, and space-intensive utilizing multiple care areas at a time when hospital capacities are already limited.^9, 10^

Second, to aid healthcare providers in assessing illness severity in COVID-19 patients, we present two predictive models of early respiratory decompensation during hospitalization: the quick COVID-19 severity index (qCSI) and a machine learning-derived COVID-19 Severity Index (CSI). These models were built on data extracted from the first four hours of care. We compare the predictive capabilities of our model to three benchmarks accessible using data in the electronic health record: the Elixhauser comorbidity index,^11^ the quick sequential organ failure assessment (qSOFA),^12, 13^ and the CURB-65 pneumonia severity score.^14^ While many clinical risk models exist, these benefit from wide clinical acceptability and relative model parsimony as they require minimal input data for calculation. The Elixhauser comorbidity index was derived to enable prediction of hospital death using administrative data.^11^ The qSOFA score was included in SEPSIS-3 guidelines and can be scored at the bedside as it includes respiratory rate, mental status, and systolic blood pressure.^12^ The CURB-65 pneumonia severity score has been well-validated for hospital disposition, but its utility in both critical illness and COVID-19 is unclear.^14, 15^

Third, we make the qCSI available to the public via a web interface at covidseverityindex.org. This web portal hosts the parsimonious model and allows for user entry of the required clinical values.

## Methods

### Study Design and setting

This was an observational study to develop a prognostic model of early respiratory decompensation in patients admitted from the emergency department with COVID-19. The healthcare system is comprised of a mix of pediatric (*n* = 1), suburban community (*n* = 6), urban community (*n* = 2), and urban academic (*n* = 1) emergency departments. Data from eight hospitals were used in the creation and internal validation of the predictive model, while data from the last site was withheld for external validation. We adhered to the Transparent Reporting of a multivariable prediction model for Individual Prognosis Or Diagnosis (TRIPOD) checklist (Supplementary Materials).^16^

### Data collection and processing

Patient demographics, summarized past medical histories, vital signs, outpatient medications, chest x-ray (CXR) reports, and laboratory results available during the ED encounter were extracted from our local Observational Medical Outcomes Partnership (OMOP) data repository and analyzed within our computational health platform.^17^ Data were collected into a research cohort using custom scripts in PySpark (version 2.4.5) that were reviewed by an independent analyst.

Non-physiologic values likely related to data entry errors for vital signs were converted to missing values based on expert-guided rules (Table S1). Laboratory values at minimum or maximum thresholds and encoded with “<” or “>” were converted to the numerical threshold value and other non-numerical values were dropped. Past medical histories were generated by using diagnoses prior to the date of admission to exclude potential future information in modeling. Outpatient medications were mapped to the First DataBank Enhanced Therapeutic Classification System.^18^ CXR reports were manually reviewed by two physicians and categorized as “no opacity”, “unilateral opacity”, or “bilateral opacities”. One hundred x-ray reports were reviewed by both physicians to determine inter-rater agreement with weighted kappa. Oxygen devices were similarly extracted from OMOP (Table S2)

### Critical respiratory illness determination

We define critical respiratory illness in the setting of COVID-19 as any COVID-19 patient meeting one of the following criteria: low-flow oxygenation ≥ 10 L/min, high-flow oxygenation, noninvasive ventilation, invasive ventilation, or death. We do not include intensive care unit admission in our composite outcome because at the start of the COVID-19 pandemic, ICU admissions were protocolized to include even minimal oxygen requirements. A subset of outcomes were manually reviewed by physician members of the institutional computational healthcare team as part of a systemwide process to standardize outcomes for COVID-19 related research.

### Inclusion and exclusion criteria

Data included visits from March 1, 2020 through April 27, 2020 as our institution’s first COVID-19 tests were ordered after March 1, 2020. This study included COVID-19 positive patients as determined by test results ordered between 14 days prior to and up to 24 hours after hospital presentation. We included delayed testing because institutional guidelines initially restricted testing within the hospital to inpatient wards. Testing for COVID-19 was performed at local and/or reference laboratories by nucleic acid detection methods using oropharyngeal (OP), nasopharyngeal (NP), or a combination OP/NP swab. We excluded patients less than 18 years of age and those who met our critical illness criteria at any point within four hours of presentation. The latter of these criteria was intended to exclude patients for whom critical illness was nearly immediately apparent to the medical provider and for whom a prediction would not be helpful. Patients who explicitly opted out of research were excluded from analysis (*n* < 5). Data were extracted greater than 24 hours after the last included patient visit so that all outcomes could be extracted from the electronic health record.

### Baseline models

We generated comparator models using Elixhauser comorbidity index, qSOFA, and CURB-65 (Supplementary Materials). ICD-10 codes from patient past medical histories were mapped to Elixhauser comorbidities and indices using H-CUP Software and Tools (*hcuppy* package, version 0.0.7).^19, 20^ qSOFA was calculated as the sum of the following findings, each of which were worth one point: GCS < 15, respiratory rate ≥ 22, and systolic blood pressure ≤ 100. CURB-65 was calculated as the sum of the following findings, each of which were worth one point: GCS < 15, BUN > 19 mg/dL respiratory rate ≥ 30, systolic blood pressure < 90 mmHg or diastolic ≤ 60 mmHg, and age ≥ 65 years. Baseline models were evaluated on the training and internal validation cohort using logistic regression on the calculated scores.

### Severity indices

Samples from eight hospitals were used in model generation and internal validation with the remaining large, urban community hospital serving as an independent dataset for external validation. All models were fit on patient demographic and clinical data collected during the first 4 hours of patient presentation. We used an ensemble technique to identify and rank potentially important predictive variables based on their occurrence across multiple selection methods: univariate regression, random forest, logistic regression with LASSO, Chi-square testing, gradient boosting information gain, and gradient boosting Shapley additive explanation (SHAP) interaction values (Supplementary Materials).^21–23^ We counted the co-occurences of the the top 30, 40, and 50 variables of each of the methods prior to selecting features for a minimal scoring model (qCSI) and machine learning model (CSI) using gradient boosting. For the qCSI, we used a point system guided by logistic regression (Supplementary Materials). The gradient-boosting CSI model was fit using the XGBoost package and hyperparameters were set using Bayesian optimization with a tree-structured Parzen estimator (Supplementary Materials).^24, 25^ All analyses were performed in Python (version 3.8.2).

### Predictive model performance

We report summary statistics of model performance in predicting the composite outcome between 4 and 24 hours of hospital arrival. We used bootstrapped logistic regression with ten-fold cross-validation to generate performance benchmarks for the Elixhauser, qSOFA, CURB-65, and qCSI models and bootstrapped gradient boosting with tenfold cross validation for the CSI model. Where necessary, data were imputed using training set median values of bootstraps. We report AU-ROC, accuracy, sensitivity and specificity at Youden’s J index, AU-PRC, Brier score, F1, and average precision (Supplementary Materials). For significance testing of AU-ROCs, we applied Welch’s t-test to average differences between permutation tests of models’ performance metrics.^26, 27^ To assess for generalizability, AU-ROCs on the external validation cohort are also presented.

### Web interface design

The qCSI was made publicly available as a web calculator at covidseverityindex.org. Nodejs, Vue, and Vuetify were used for the website front-end.

### Patient and public involvement

This was a retrospective observational cohort study and no patients were directly involved in the study design, setting the research questions, or the outcome measures. No patients were asked to advise on interpretation or presentation of results. This study was approved by our local institutional review board (IRB# 2000027747). Dissemination to study participants is not applicable.

### Results

Between March 1, 2020 and April 27, 2020, there were a total of 1,792 admissions for COVID-19 patients. Of these, 620 patients (35%) were excluded by meeting critical respiratory illness endpoints within 4 hours of presentation or by age criteria. Of the included patients, 144 (12.3%) had respiratory decompensation within the first 24 hours of hospitalization: 101 (8.6%) requiring >10 L/min oxygen flow, 112 (9.6%) on a high-flow device (Table S2), 4 (0.3%) on non-invasive ventilation, 10 (0.8%) with invasive ventilation, and 1 (0.01%) death. 59 (5%) of patients were admitted to the ICU with the 4 to 24 hour time period. Population characteristics including demographics and comorbidities for the study are shown in Table 1. Study patient flow is shown in Figure 1.

**Table 1:**
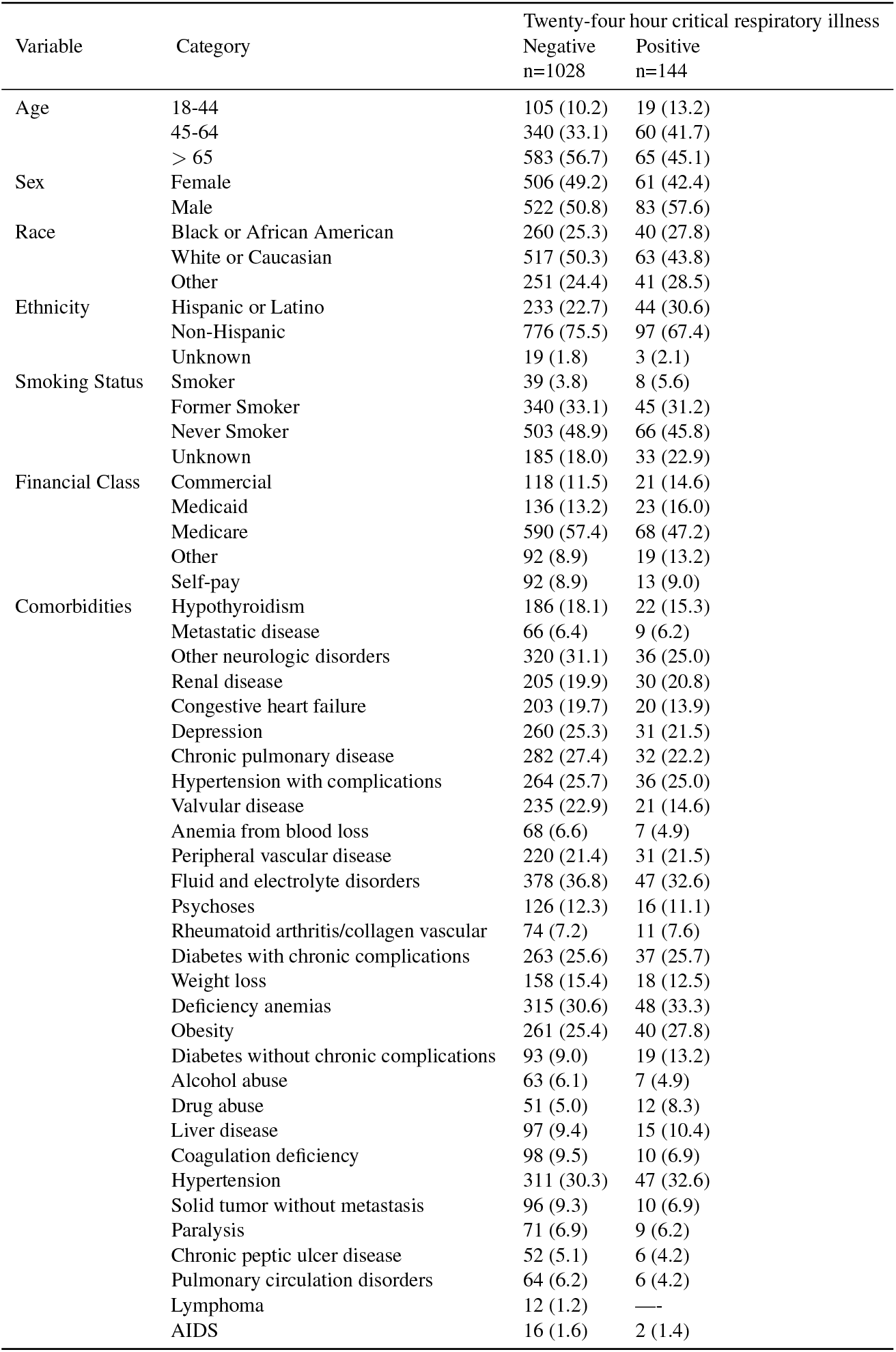
Characteristics of COVID-19 positive admitted patients stratified by primary outcome

**Figure 1:**
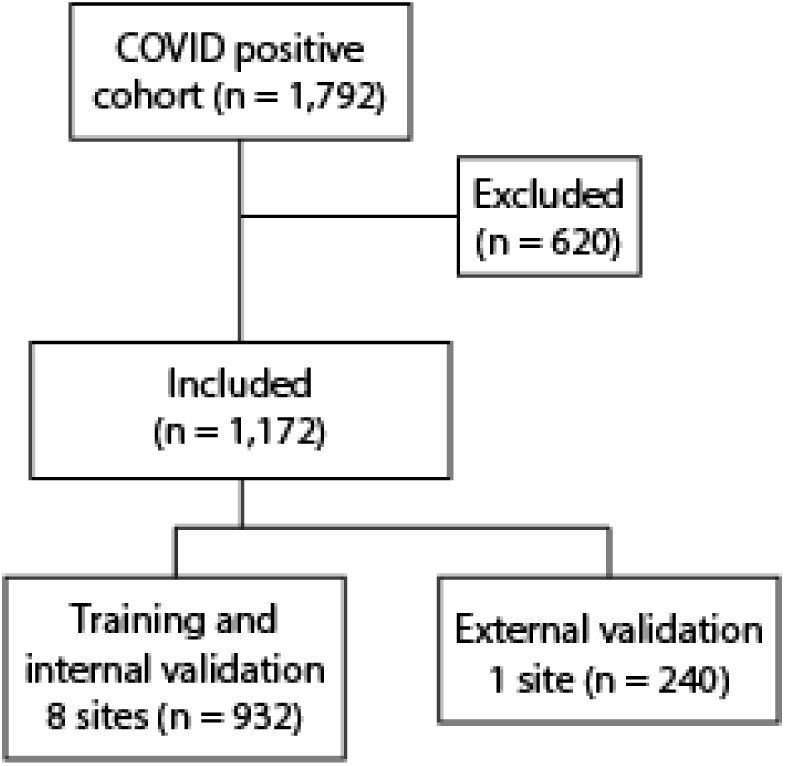
Study flow diagram

### Identification of predictive factors for critical illness

Our full dataset included 713 patient variables available during the first four hours of the patient encounters (Table S3). These included demographics, vital signs, laboratory values, comorbidities, chief complaints, outpatient medications, tobacco use histories, and CXR. Radiologist evaluated CXRs were classified into three categories with strong inter-rater agreement (κ = 0.81). Associations between CXR findings and outcomes are shown in Table S4. We preferentially selected variables available at bedside for creation of the qCSI. Our ensemble approach identified three bedside variables as consistently important across the variable selection models: nasal cannula requirement, minimum recorded pulse oximetry, and respiratory rate (Figure S1). These three features appeared in at least five of the six variable selection methods.

### qCSI and CSI variable weights

We divided each of these three clinical variables into value ranges using clinical experience and used logistic regression to create weights for the qCSI scoring system (Table 2). Normal physiology was used as the baseline category, and the logistic regression odds ratios were offset to assign normal clinical parameters zero points in the qCSI (Supplementary Materials).

**Table 2:**
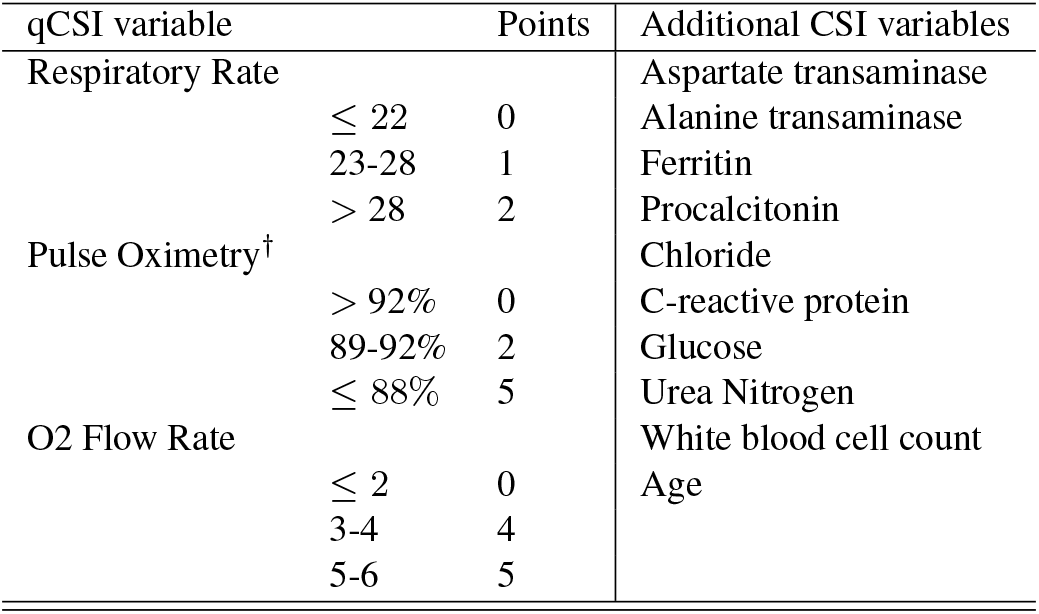
qCSI and CSI model variables. ^†^ Pulse oximetry represents the lowest value recorded during the first four hours of the patient encounter.

We identified an additional twelve features from the predictive factor analysis for use in a machine learning model (CSI) with gradient boosting (Table 2, Figure S1). These variables were selected by balancing the goals of model parsimony, minimizing highly correlated features (i.e., various summaries of vital signs), and predictive performance. We used SHAP methods to understand the importance of various clinical variables in the CSI (Figure 2).^23, 28–30^ SHAP values are an extension of the game-theoretic Shapley values that seek to describe variable impacts on model output, as defined as the contribution of a specific variable to the prediction itself.^28^ The key advantage of the related SHAP values is that they add interpretability to complex models like gradient boosting, which otherwise provide opaque outputs. SHAP values are dimensionless and represent the log-odds of the marginal contribution a variable makes on a single prediction. In the case of our gradient boosting CSI model, we employ an isotonic regression step for model calibration, so the SHAP values reflect a relative weighting of contributions.^31^

**Figure 2:**
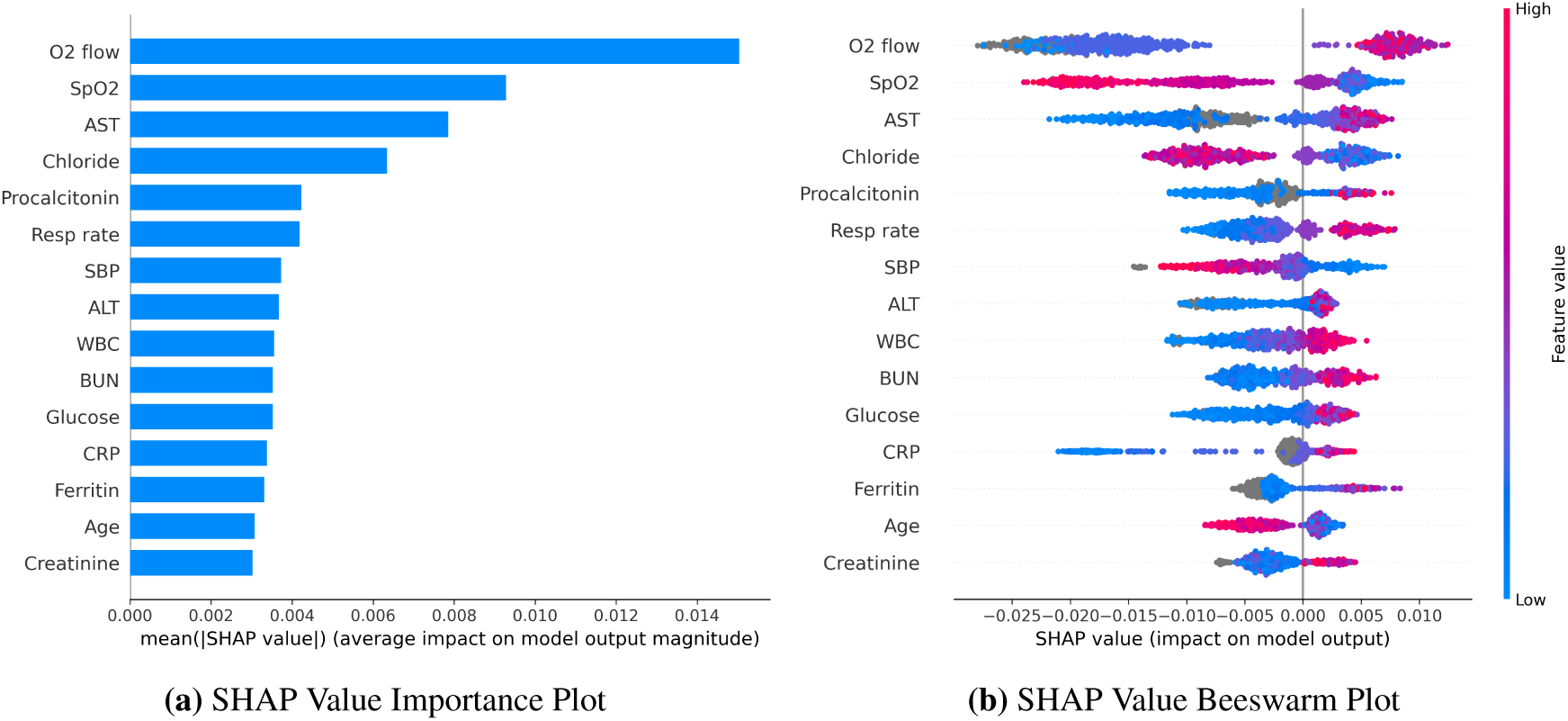
SHAP variable importance and beeswarm plots. (a) Mean absolute SHAP values suggest a rank order for variable importance in the CSI. (b) In the beeswarm plot, each point corresponds to an individual person in the study. The points position on the x-axis shows the impact that feature has on the model’s prediction for a given patient. Color corresponds to relative variable value.

The rank-order of average absolute SHAP values across all variables in a model suggests the most important variables in assigning modeled risk. For the CSI these were flow rate by nasal cannula, followed by lowest documented pulse oximetry, and aspartate aminotransferase (AST) (Figure 2a). Consistent with prior studies, we also observed utility of inflammatory markers, ferritin, procalcitonin, and CRP. We then explored how ranges of individual feature values affected model output (Figure 2b). For example, low oxygen flow rates (blue) are protective as indicated by negative SHAP values, as are high pulse oximetry values (red). To better investigate clinical variable effects on predicted patient risk, we generated individual variable SHAP value plots (Figure 3). Age displayed a nearly binary risk distribution with an inflection point between 60 and 70 years of age (Figure 3a). Younger patients displayed a higher risk of 24 hour critical illness than did older patients. We also observed that elevated AST, alanine aminotransferase (ALT), and ferritin were associated with elevated model risk, but the SHAP values reached their asymptotes well before the maximum value for each of these features (Figure 3b-d). AST and ALT SHAP values reached their maximum within normal or slightly elevated ranges for these laboratory tests. The inflection point in risk attributable to ferritin levels, however, was close to 1000 ng/mL, above institutional normal range for this test (30-400 ng/mL).

**Figure 3:**
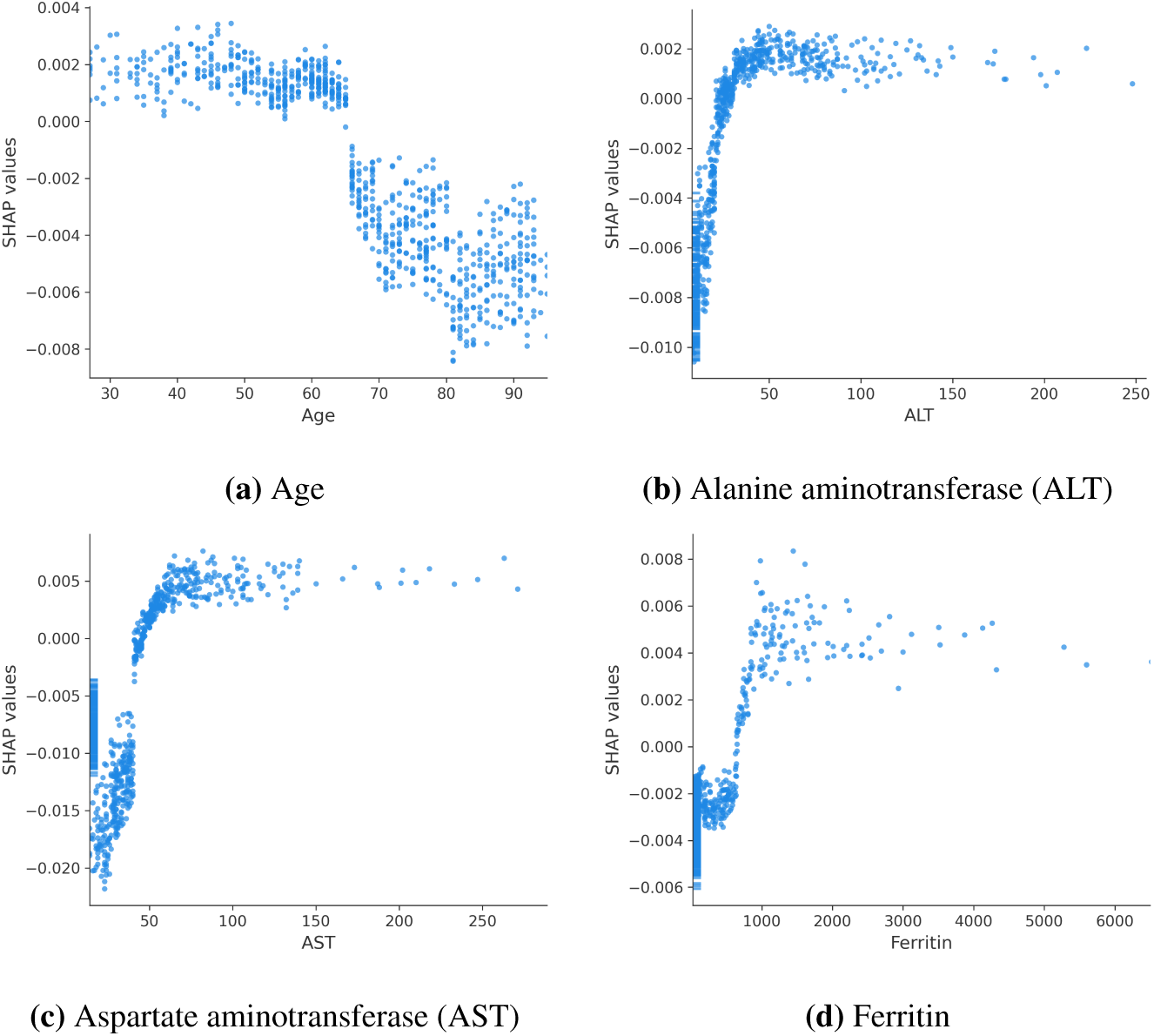
SHAP value plots for (a) age, (b) alanine aminotransferase, and (c) aspartate aminotransferase, and (d) ferritin. Scatter plots show the effects of variable values (x-axis) on the model predictions as captured by SHAP values (y-axis).

### qCSI and CSI performance

The qCSI (0.89 [0.84, 0.95]) and CSI (0.92 [0.86, 0.97]) models outperformed the comparator models with the CSI best by the AU-ROC metric overall (Table 3). Among the evaluated benchmark models, qSOFA (AU-ROC, 95% CI; 0.76 [0.69-0.85]) was better than either Elixhauser (0.70 [0.62-0.80]) or CURB-65 (0.66 [0.58-0.77]) in predicting the composite endpoint (*p* < 0.05 for all comparisons).

**Table 3:**
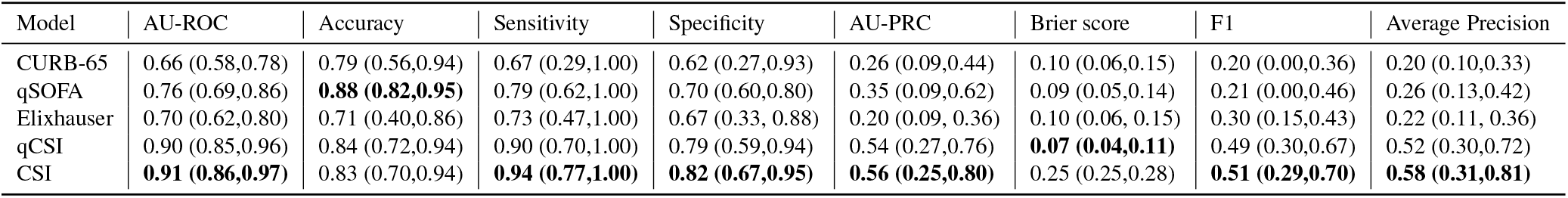
Performance Characteristics for CSI, qCSI, and comparison models on cross-validation. The best performance for a given metric is shown with bold text.

### External validation

We then tested the predictive performance of qCSI and CSI on the external validation cohort in order to test their generalizability, finding AU-ROC of 0.82 and 0.76, respectively. The baseline models had worse performance on external validation (qSOFA 0.59, CURB-65 0.50, Elixhauser 0.61). We also tested the calibration of the qCSI and CSI scores by assigning all patients in the external cohort each of the scores and comparing them to known outcomes (Figure 4, Figure S2-3).^32^ These calibration curves suggest that outcome rates increased with qCSI and CSI scores.

**Figure 4:**
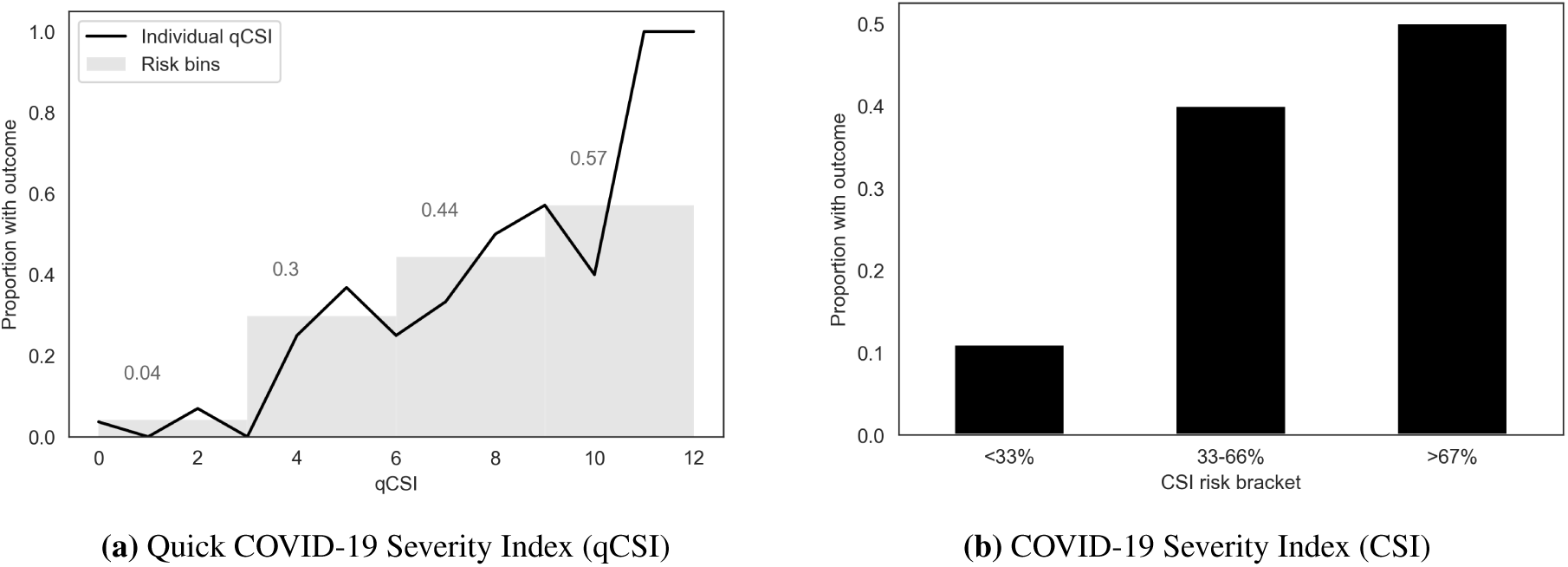
Calibration of qCSI and CSI on external validation dataset. (a) Each patient in the validation cohort was assigned a score by qCSI, and the percentage who had a critical respiratory illness outcome were plotted with a line plot. Patients were then grouped into risk bins by qCSI intervals (0-3, 4-6, 7-9, 10-12) and the percentage of patients in each group with the outcome is indicated in the bar plot. (b) Each patient in the validation cohort was assigned a CSI, a percent risk from 0 to 100% using gradient boosting and isotonic regression. The percent of patients with CSI scores of 0-33%, 33-66%, and 66-100% who experienced critical respiratory illness at 24 hours are shown.

### Web application

The qCSI is available at covidseverityindex.org. The qCSI calculator includes selection boxes for each of the three variables which are summed to generate a score and prediction as estimated using the external validation cohort.

## Discussion

Consistent with clinical observations, we noted a significant rate of progression to critical respiratory illness within the first 24 hours of hospitalization in COVID-19 patients. We used six parallel approaches to identify a subset of variables for the final qCSI and CSI models. The qCSI ultimately requires only three variables, all of which are accessible at the bedside.

We propose that a qCSI score of 3 or less be considered low-likelihood for 24 hour respiratory critical illness with a mean outcome rate of 4% in the external validation cohort (Figure 4). This score is achievable under the following patient conditions: respiratory rate ≤ 28, minimum pulse oximetry reading of ≥ 89%, and oxygen flow rate of ≤ 2L. We note, however, that few patients in the validation cohort had qCSI of 3 (SpO2 of 89-92% and respiratory rates of 23-28 with ≤ 2 L/min oxygen requirement)(Figure S2). Patients with a qCSI of 4-6 had a 30% rate of decompensation, while the 7-9 group had a 44% rate, and the 10-12 group a 57% rate. We noted limited numbers of patients in the upper risk groups - larger study sizes will be required to clarify their clinical significance.

While statistically significant, the modest increases in performance of the CSI as compared to the qCSI suggest that the more parsimonious qCSI is likely preferable for rapid implementation. Comparison between qCSI and CSI on the external validation cohort offers one evaluation of potential generalizability, but further studies will be required. The CSI, however, offers opportunities to perform further analysis of potential COVID-19 prognostic factors. In alignment with current hypotheses about COVID-19 severity, we note that multiple variable selection techniques identified inflammatory markers including CRP and ferritin as potentially important predictors. More striking however, was the importance of AST and ALT in CSI predictions as calculated with SHAP values.^33, 34^ Lower age had higher SHAP values, suggesting potential bias in the admitted patient cohort - young, admitted patients may be more acutely ill than older admitted COVID-19 patients. Interestingly, the transition point where the SHAP value analysis identified model risk associated with liver chemistries was at the high end of normal, consistent with previous observations that noted that normal to mild liver dysfunction among COVID-19 patients. We hypothesize that the asymptotic quality of the investigated variables with respect to CSI risk contributions reflects our moderate study size. We expect that scaling CSI training to larger cohorts will further elucidate the impacts of more extreme lab values. While our dataset included host risk factors including smoking history, obesity, and BMI, these did not appear to play a prominent role in predicting deterioration. Here, we recognize two important considerations: first, that predictive factors may not be mechanistic or causative factors in disease, and second that these factors may be related to disease severity without providing predictive value for 24 hour decompensation.

We include CXRs for 1,170 visits in this cohort. CXR are of significant clinical interest as previous studies have shown high rates of ground glass opacity and consolidation.^35^ Chest CT may have superior utility for COVID-19 investigation, but are not not being widely performed at our institutions as part of risk stratification or prognostic evaluation.^36, 37^ CXR reports were classified based on containing bilateral, unilateral, or no opacities or consolidations. We found high inter-rater agreement in this coding, but CXR were not consistently identified by our variable selection models. A majority of patients were coded as having bilateral consolidations, limiting the specificity of the findings. Further studies using natural language processing of radiology reports or direct analysis of CXR with tools like convolutional neural networks will provide more evidence regarding utility of these studies in COVID-19 prognostication.^38^ Furthermore, we do not consider other applications of CXR including the identification of other pulmonary findings like diagnosis of bacterial pneumonia.

The Elixhauser comorbidity index, qSOFA, and CURB-65 baseline models provided the opportunity to test well-known risk stratification and prognostication tools with a COVID-19 cohort. These tools were selected, in part, for their familiarity within the medical community, and because each has been proposed as having potential utility within the COVID-19 epidemic. We note the relatively limited predictive performance of these metrics, while simultaneously recognizing that none were designed to address the clinical question addressed here. In particular, the CURB-65 pneumonia severity score may still have utility in determining patient disposition with respect to discharge or hospitalization.

Future studies will be required to expand on this work in a number of ways. First, prospective, multi-site validation is required for the qCSI. The CSI may lend itself to a “living” model framework where the addition of new features, weights, and outcomes will improve its predictive capability.^8, 39^ We hypothesize that the CSI will continue to improve as compared to the qCSI as more patient observations are included. Second, we expect related models to be extended to patient admission decisions as well as continuous hospital monitoring.^40–42^ The qCSI does not separate patients without any nasal cannula requirement from those with even a minimal oxygen requirement. We expect that future models for safe discharge of COVID-19 patients will more strongly weigh even low oxygen requirements as local practice patterns may likely necessitate admission of any patient on exogenous oxygen.

Patient prognosis has important ramifications in terms of resource utilization, hospital placement, and patient shared decision-making. We additionally note the role of respiratory parameters in selecting patients for therapeutic interventions. An early proof-of-concept study for the viral RNA polymerase inhibitor Remdesivir, which has *in vitro* activity against SARS-CoV-2, included patients with pulse oximetry of ≤ 94% on ambient air or who had any oxygen requirement.^43^ There is a large ongoing clinical trial that uses similar inclusion criteria (ClinicalTrials.gov Identifier: NCT04292899). A 237 patient Chinese trial of the same drug was stopped early after no further eligible patients were available for enrollment.^44^ This study included patients with confirmed COVID-19 infection by RT-PCR, pneumonia on imaging, oxygen saturation of ≤ 94% on ambient air, or a partial pressure to fractional inspired oxygen ratio of 300 mm Hg or less. Improved pragmatic, prognostic tools like the qCSI may offer a route to expanded inclusion criteria for ongoing trials or for early identification of patients who might potentially benefit from therapeutics.

### Limitations

The data in this study were observational data provided from a single health system and so may not be generalizable based on local testing and admissions practices. Our data were extracted from an electronic health record, which is associated with known limitations including propagation of old or incomplete data. Similarly, there are important markers of oxygenation which were out of the scope of our study, including alveolar-arterial gradients.

Retrospective observational studies lack control of variables so prospective studies will be required to assess validity of the presented models and the specificity of the features we identify as important to COVID-19 progression. Assumptions were made in data processing where noted in the methods, which introduce biases into our results. Chest x-ray interpretation was done manually using radiology reports, but without reviewing the radiography, which introduces subjectivity as reflected in the inter-rater agreement metric. Most significant, however, is that management of COVID-19 is evolving, so it may be possible that future clinical decisions may not match those standards used in the reported clinical settings.

## Conclusions

The qCSI robustly predicts clinical respiratory decompensation in COVID-19 patients using pulse oximetry, respiratory rate, and nasal cannula flow rate. The CSI highlights the associations of a number of variables including liver chemistries and inflammatory markers with patient risk. Prospective, multi-site validation will be required to better assess the generalizability of these models. The qCSI is available at covidseverityindex.org.

## Data Availability

Due to patient privacy concerns, the data in this study cannot be made publicly available.

## Contribution details

ADH, NGR, RAT designed the project. NGR, HPY, WLS, RAT extracted and processed the data. ADH, NGR, and RAT created the models with guidance from FPW and DvD. SS designed the web interface. All authors contributed substantially to manuscript revisions. RAT is the guarantor.

## Data sharing statement

Due to patient privacy, the data are not available for distribution.

## Ethical approval

This study was approved by our local institutional review board (IRB# 2000027747).

## Patient consent

N/A.

## Data sharing

No additional data available.

## Dissemination to participants and related patient and public communities

This was a retrospective observational cohort study and no patients were directly involved in the study design, setting the research questions, or the outcome measures. No patients were asked to advise on interpretation or presentation of results.

